# FarmApp: a new cognitive assessment method for young people with intellectual disability

**DOI:** 10.1101/2020.12.02.20242677

**Authors:** Diandra Brkić, Elise Ng-Cordell, Sinéad O’Brien, Jessica Martin, Gaia Scerif, Duncan Astle, Kate Baker

## Abstract

**Background:** A major challenge when investigating intellectual disability (ID) is the selection of assessment tools that are sensitive to cognitive diversity within the ID population. This study introduces a new touchscreen-based method, FarmApp, which aims to measure competence in relatively low-level cognitive processes (selective attention, short-term visuospatial memory, longer-term recognition memory) which contribute to complex aspects of learning and behaviour.

**Methods:** Here we describe the FarmApp design, testing and analysis procedures. We report the feasibility and validity of the method, and demonstrate its utility for measuring change over time, and for comparing groups defined by aetiology.

**Results:** We found that FarmApp can be completed by a higher proportion of young people with ID than traditional psychometric tests. FarmApp performance correlates with standardised neuropsychological tests of attention and working memory, and with questionnaire measures of ADHD-relevant behavioural difficulties. Individual performance slopes over a two-week period correlate with general ability and behavioural difficulties, indicating that FarmApp is sensitive to meaningful dynamic variation in cognitive performance. Finally, we compared the FarmApp performance of two groups of young people with ID, defined by the physiological function of ID-associated genetic variants (functional network groups: chromatin-related and synaptic-related), and found that groups differ on attention parameters but not on memory ones.

**Conclusion:** FarmApp is a feasible, valid and useful alternative to traditional neuropsychological tests. It can increase access to cognitive assessment for individuals with ID. It adds the opportunity to monitor variation in performance over time and determine capacity to acquire task competence in addition to baseline ability. Our comparison between functional network groups supports the proposal that cognitive processes contributing to ID are differentially influenced by specific genetic aetiologies. In summary, we introduce a new tool for cognitive assessment in ID, with the potential for multiple future applications in clinical practice and research.

## Introduction

Intellectual disability (ID) is defined by significant limitations to cognitive and adaptive functions, with onset during childhood (1). There is wide variation in the types and severity of cognitive difficulties affecting individuals with ID, in turn influencing variation in educational attainment, independent living skills, social interactions, and emotional characteristics (2). Moreover, the same degree of global impairment (mild, moderate or severe ID) and same behavioural characteristics (e.g. hyperactivity or anxiety) can arise from different underlying cognitive mechanisms (3). For example, sensory functions, motor control or memory limitations can each be the primary mechanism contributing to communication difficulties for an individual with ID (4). A better understanding of the low-level cognitive processes contributing to ID, cognitive diversity within the ID population, and the relationship between causal factors and cognitive mechanisms could be of practical use, and provide an important bridge between the biological origins of ID and complex developmental impairments. In view of rapid progress in identifying genetic aetiologies of ID, it should become possible to test whether the cause of an individual’s ID influences learning via specific cognitive processes (5).

A longstanding challenge in this field has been the lack of assessment measures sensitive to cognitive diversity within the ID population. The use of standardised intelligence tests or neuropsychological measures, whilst providing uniform benchmarking, poses several limitations (6). Firstly, individuals with ID may have difficulties understanding task goals, attending to task instructions, or processing complex task stimuli, leading to underestimation of their actual abilities within the domain under assessment. Secondly, strict administration procedures, for example time limits or required number of items that must be completed for scoring, means that an individual may not achieve recognition for their performance. Furthermore, standardization samples usually include very few subjects with ID and associated syndromes, resulting in limited sensitivity within range of impairment for individuals with ID, and frequent floor effects. There have been significant attempts to overcome these limitations (7-9), however existing methods rely on a one-off assessment by an unfamiliar examiner, irrespective of an individual’s variable concentration, motivation and social confidence, all of which may influence capacity to attempt a novel task and to improve at a task if practise is permitted (10).

We have designed an accessible touchscreen-based method – FarmApp – that aims to measure competence in relatively low-level cognitive processes. For example stimulus-response accuracy and speed, short-term memory capacity, and longer-term recognition, which contribute to diverse aspects of everyday function. FarmApp is an intuitive cognitive assessment platform, not relying on verbal instructions, using age and developmental-stage appropriate materials and a game-like structure that are motivating and enjoyable for participants. Influenced in part by cognitive training methods, which often involve adaptive cognitive tasks, FarmApp introduces a number of elements of flexibility to improve access and maximise the opportunity for each participant to demonstrate their abilities (11). Participation can be supervised by a researcher, a parent / carer, or can be independently controlled by a participant (12). The computerised testing schedule is programmed in an adaptive fashion, to maintain motivation and obtain an estimate of “best possible” performance (11, 13). The app is designed to be used repeatedly over a period of days or weeks, with no fixed requirement or limit to number of attempts, to monitor changes in a participant’s performance via remote data upload. The selection of tasks was informed by our core research hypotheses that distinct cognitive processes can contribute to complex learning impairments reflecting different aetiologies of ID, and that dynamic change in cognitive performance may be more informative than fixed deficits (14, 15). Specifically, we sought to extend our previous findings that genetic disorders directly influencing synaptic physiology impact on attention and cognitive control, contributing to social and behavioural difficulties amongst this group (16).

In the current paper, we describe the FarmApp design, testing and analysis procedures, and we address the following issues and questions:

1. Feasibility - Can FarmApp achieve its objectives in populations that typically struggle to complete traditional psychometric assessments? Does longer-term testing at home improve access to assessment?
2. Validity - Can FarmApp be used to characterise underlying cognitive processes which contribute to learning, adaptive functions and everyday behaviour? To establish this, we asked whether FarmApp tasks are comparable to standardised neuropsychological measures, and whether FarmApp performance relates to parent-reported measures of everyday behaviours.
3. Utility of assessing performance over time - Can FarmApp incorporate assessment of change in performance over time to capture dynamic capacities beyond baseline constraints? Does change in task performance over time relate to an individuals’ global adaptive function and behavioural characteristics?
4. Effectiveness of the method to link causal factors and cognitive mechanisms - As proof of principle, we examined whether FarmApp task performance and dynamic change in attention task performance are associated with ID-associated genetic variants. We applied the functional network phenotyping approach whereby variants are grouped according to gene physiological function (16).

## Method

### FarmApp structure

FarmApp consists of three independent tasks, or games, which are described below. The three games share a similar overarching structure. These can be accessed in one of three “modes”. In RA mode, the research assistant introduces the task to the participant, and guides the participant as they progress through the game. This is the mode that most closely resembles existing experimenter driven assessments, albeit with ID friendly modifications [e.g. (8, 9)]. However, and uniquely, once the participant has completed the games in RA mode, FarmApp is switched into Child mode, in which participants are able to play the games freely and at their own pace. Additionally, there is an option to enter Adult-mode, where adults (usually parents) can intervene and guide their child through the rules and procedure of each game.

Within each mode, the games are further divided into “phases”. RA mode consists of an initial “training” phase, in which the rules and requirements of the game are explained alongside a simple narrative (see supplementary material). The training phase can be repeated until the participant understands the game enough to proceed to the next phase, having completed one block per game. The next phase is “warm-up”, allowing the participant to attempt several trials with reduced supervision and prompting from the RA. After the warm-up phase has been completed, the game progresses into the “baseline” phase, in which the participant completes full-length blocks of each game. Performance at baseline is used to establish a starting point for play during the final “adaptive” phase – in which task difficulty increases or decreases according to each participant’s performance. In Child mode, there are no training and warm-up phases; only baseline and adaptive phases. Each time one of the games is opened, a new “run” for that game is initiated. A run is complete when the participant has progressed through each phase of the game. Our motivation behind this structure was to enable adaptive testing for as many participants as possible. The structure and design, including the three modes (RA, child, parent) and four phases (training, warm-up, baseline, and adaptive), were finalised after piloting with children (with and without ID).

### Administration procedures

For the current study, all participants were initially assessed in a quiet room in their home or at school. A research assistant explained the objectives and functions of FarmApp to participants and their parents and carers, before introducing the three games within the app to the participant and guiding them through RA mode. After this, FarmApp entered Child mode, and the tablet was left with the family for a period of two weeks. Parents were asked to encourage their child to use the app independently for about 20 minutes every day. During this period, the study team also provided technical support via email and telephone to families as needed, and were able to monitor task activity remotely. Tablets were returned via courier service at the end of the two weeks.

### FarmApp Tasks

A brief summary of each FarmApp game is provided below. For detailed descriptions and a complete list of variable definitions, please see supplementary material.

#### Sheep Game

A go/no-go task in which participants are asked to “catch” as many sheep as possible, while avoiding any pigs. Images of sheep or pigs are visually presented one at a time in the bottom left corner of the screen. In order to catch a sheep, the participant must tap on the sheep while it is presented on the screen. Audio feedback is provided for a correct response (“baa” for a captured sheep, and “oink” for an untouched pig). Initially during training, warm-up, and baseline phases, the stimuli are presented for a set duration (“stimulus duration”) of 3000ms. During the later adaptive phases, stimulus duration is adjusted according to participants’ accuracy at no-go trials: after a correctly inhibited response to a pig, stimulus duration decreases by 20%; after an incorrectly uninhibited response to a pig, stimulus duration increases by 20%. In order to keep the games engaging and feasible, minimum and maximum stimulus durations were capped at 1000ms and 8000ms, respectively. We recorded participants’ accuracy (“overall accuracy”), which we further broke down into accuracy when capturing sheep (“go accuracy”) and avoiding pigs (“no-go accuracy”). We also recorded participants’ response time (“RT”) when capturing sheep during blocks containing only sheep in the baseline phase and in the adaptive one (“go-only RT”). Finally, during the adaptive phase, we recorded “adaptive RT” averaged across mixed trials (go-only and no-go), and the duration of no-go stimulus presentation (“stimulus duration”), as it changed according to participants’ accuracy on no-go trials, as a metric for change in performance over time.

#### Chicken Game

a Corsi block test where the stimuli are chickens popping out of hutches. The participants are asked to tap on the hutches in the same order that the chickens appeared. The participant has, again, audio feedback for a correct sequence recalled and by seeing a last chicken in the sequence lay an egg. The eggs (rewards) accumulate over multiple runs, appearing in a haystack. During the Adaptive phase, number of stimuli presented adjusts to the participant’s accuracy on the previous block. Variables of interest include maximum span reached and overall accuracy. In addition, in order to account for variability within the same level (three, four, or five hutches) of difficulty, a weighted score is computed, taking into the account the proportion of correct number of trials at the span level.

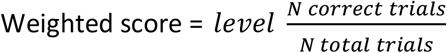

#### Memory game

assessment of recognition for items presented in the Chicken and Sheep games vs novel distractor items from matched stimulus sets. The target stimuli (10 trials or pairs per block) are generated from sheep or chicken stimuli seen during previous Chicken or Sheep games. Forced-choice pairs are presented, and the participant is asked to tap on the stimulus (chicken or sheep) they have seen before. In this task, audio feedback is played to reward a response, but unrelated to accuracy. Here, the variable of interest is accuracy, and there is no adaptive element.

### Participants and recruitment

This study involved participants from two research cohorts based at MRC Cognition and Brain Sciences Unit, University of Cambridge. Overall, there were 88 participants recruited for this study, where 61 were included in at least one of the analyses. Of these, 18 children participated from the CALM (Centre for Attention, Learning and Memory) cohort. Core inclusion criteria for the CALM cohort were aged 5-18 years and native English speakers, with cognitive and/or learning problems identified by a health or educational professional. A full description of recruitment procedures and cohort characteristics is provided in the CALM protocol paper (17). In addition, 43 individuals participated from the BINGO (Brain and Behaviour in Intellectual Disability of Genetic Origin) cohort. Core inclusion criteria for the BINGO cohort were age three and above, diagnosed with a neurodevelopmental disorder of known genetic origin (see supplementary material for the list of genetic disorders). Participants were recruited to BINGO after receiving their genetic diagnosis, with the assistance of Regional Genetics Centres and family support groups. Demographic information for participants from both cohorts in the FarmApp study is present in Table 1, together with breakdown of participant numbers involved in each subsequent analysis.

**Table 1.** Demographic summary.

|  | CALM<br>PARTICIPANTS | BINGO<br>PARTICIPANTS | TOTAL<br>SAMPLE |
| --- | --- | --- | --- |
| Number | 18 | 43 | 61 |
| Age / months<br>Mean (range, SD) | 120 (75-163; SD=25) | 171 (69-315; SD=66) | 156 (69-315; SD=62) |
| Gender (F:M) | 9:9 | 24:19 | 33:28 |
| Vineland ABC<br>Mean (range, SD) | 90 (59-127; SD=24) | 57 (20-96; SD=14) | 66 (20-127; SD=21) |

### Concurrent Psychometric Measures

Parents / carers completed the Vineland Adaptive Behaviour Scales – second edition (VABS2) as a measure of global adaptive function (18), and the Conners Parent Rating Scales – Third edition, short form (CPRS) as an assessment of ADHD-relevant behaviours (19).

Participants from CALM completed the Wechsler Abbreviated Scale Intelligence, second edition (WASI-II) (20). Participants from BINGO attempted either the WASI-II or Leiter international performance scale-revised (21).

To validate the Sheep FarmApp task, we used raw scores from subtests of The Test of Everyday Attention for Children 2 (TEA-Ch2) which evaluate sustained attention, switching and mental flexibility (22). Children from the CALM cohort completed the Simple Reaction Time (SRT) test. Children aged eight and above completed the Reds, Blues, Bags and Shoes (RBBS) set-switching task.

To validate the Chicken FarmApp task, we used raw scores from the Dot Matrix subtest of the Automated Working Memory Assessment (AWMA), as a measure of visuospatial short term memory (23, 24).

### Statistical analysis

To assess feasibility, we considered the proportion of individuals within the cohort who provided analysable data in RA mode and in Child mode for each game (Sheep, Chicken, and Memory), in relation to proportion of those able to complete standardised IQ testing (WASI or Leiter). Specifically, participants were included in analysis of RA mode data if they satisfied the criteria of minimum five no-go adaptive trials played in the Sheep game, at least one level of hutches completed in the chicken game, and minimum of one run for the memory game. For Child mode, we applied the same inclusion criteria as for the RA-mode.

To assess validity, we asked whether RA mode chicken and Sheep game performance correlated with psychometric test performance (TeaCh2 and AWMA), using non-parametric ranked (Spearman’s) correlations, controlling for both age and general ability (Vineland ABC). Correlations were performed in SPSS, and correlation matrices were visualised using the R *corrplot* statistical toolbox (25, 26).

To assess change in performance over time and its relation to behavioural and cognitive factors in participants with ID, we applied linear mixed effect models (LMMs) to study individual learning trajectories, and the effect of behavioural variables on their slopes. These analyses were performed in R, using the *lmer* toolbox (27). We chose to apply LMMs over a simple general linear model, because of their proven ability to deal with incomplete datasets (28).

Finally, we explored how genetic diagnosis influenced the performance at the FarmApp games. We compared Chicken, Sheep, and Memory game performance in Child mode, between two functional genetic groups: chromatin-related variants versus synaptic-related variants (see supplementary material for gene lists and participant numbers). We allocated single genes to two functional network groups (FNG): genes in the chromatin group are chromatin structural modifiers (e.g. components and regulators of the SWI/SNF chromatin remodelling complex), and genes in the synaptic group encompass direct and indirect modifiers of synaptic physiology. The two FNG’s FarmApp performance were compared via ANCOVAs, controlling for general ability (Vineland ABC) and age, in R.

## Results

### 1. Feasibility

Of the 88 participants from the two cohorts (BINGO and CALM), 56.3% were able to provide analysable data on at least one of the FarmApp games in RA mode. This increased to 69.3% providing analysable data on at least one FarmApp game in Child mode. This compares to 50% of the cohort who were able to complete a standardised IQ test (WASI-II for CALM, WASI-II or Leiter-R for BINGO). Table 2 provides a breakdown of participation across the three games and two modes of testing.

**Table 2:** Participants completing each game in RA mode and Child mode.

|  | Chicken game |  | Sheep game |  | Memory game |  |
| --- | --- | --- | --- | --- | --- | --- |
| **RA mode** |  |  |  |  |  |  |
| N (%) participants meeting inclusion criteria for analysis | 45 (56.3%) |  | 42 (47.7%) |  | 7 (8.75%) |  |
| Completers vs non-completers age / months | 155.34 | 142.61 | 151.88 | 146.07 | 150.29 | 148.71 |
| Completers vs non-completers general ability / VABC | **72.73** | **52.80** | **72.54** | **54.09** | 66 | 62.61 |
| **Child mode** |  |  |  |  |  |  |
| N (%) participants meeting inclusion criteria for analysis | 61 (69.3%) |  | 61 (69.3%) |  | 26 (29.5%) |  |
| Completers vs non-completers age | **157.68** | **128.42** | 150.98 | 143.88 | 156.15 | 145.67 |
| Completers vs non-completers VABC | **66.62** | **54.7** | **67.32** | **52.65** | 68.85 | 60.30 |

Highlighted in bold significant differences between the groups (t-test, $p < 0.05$ uncorrected for multiple comparisons).

### 2a. Validation against standardised tests

We assessed the relationships between performance on the Sheep and Chicken games (RA mode, adaptive phase) and standardised test performance, in the CALM sample only. The rationale for assessing validity in the CALM sample was that standardised neuropsychological tests are accessible to these (higher IQ) participants, but are not universally accessible to the BINGO sample. FarmApp RA mode data was selected for validation because of similar administration procedure to TEA-Ch2 (one-off, with researcher present). All trials for each FarmApp game were collapsed across blocks and runs to obtain average accuracy and response time (RT) measures. For this analysis, we carried out pairwise (Spearman Rho) correlations for each game separately, co-varying for age and general ability (Vineland ABC). Correlation matrices are shown in Figure 2 (panels a and b). Focusing on comparable measures in the Sheep game, go-only RT and TEA-Ch2 SRT raw scores were not correlated (*r*_*s*_=.244, p>.05, N=13); mixed (go and no-go) blocks RT positively correlated with TEA-Ch2 RBBS raw RT (*r*_*s*_=.894, p<.05, N=10) when controlling for age and Vineland ABC. In the Chicken game, adaptive weighted accuracy positively correlated with AWMA Dot matrix raw score when correcting for age and Vineland ABC (*r*_*s*_=.509, p<.05, N=14).

**Figure 1.**
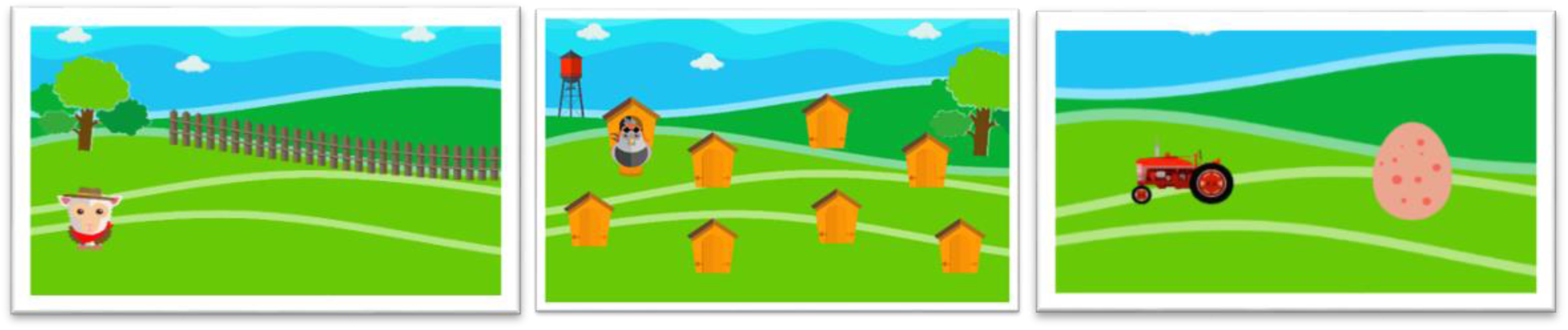
FarmApp game frames. From left to right, Sheep, Chicken and Memory game frames.

**Figure 2.**
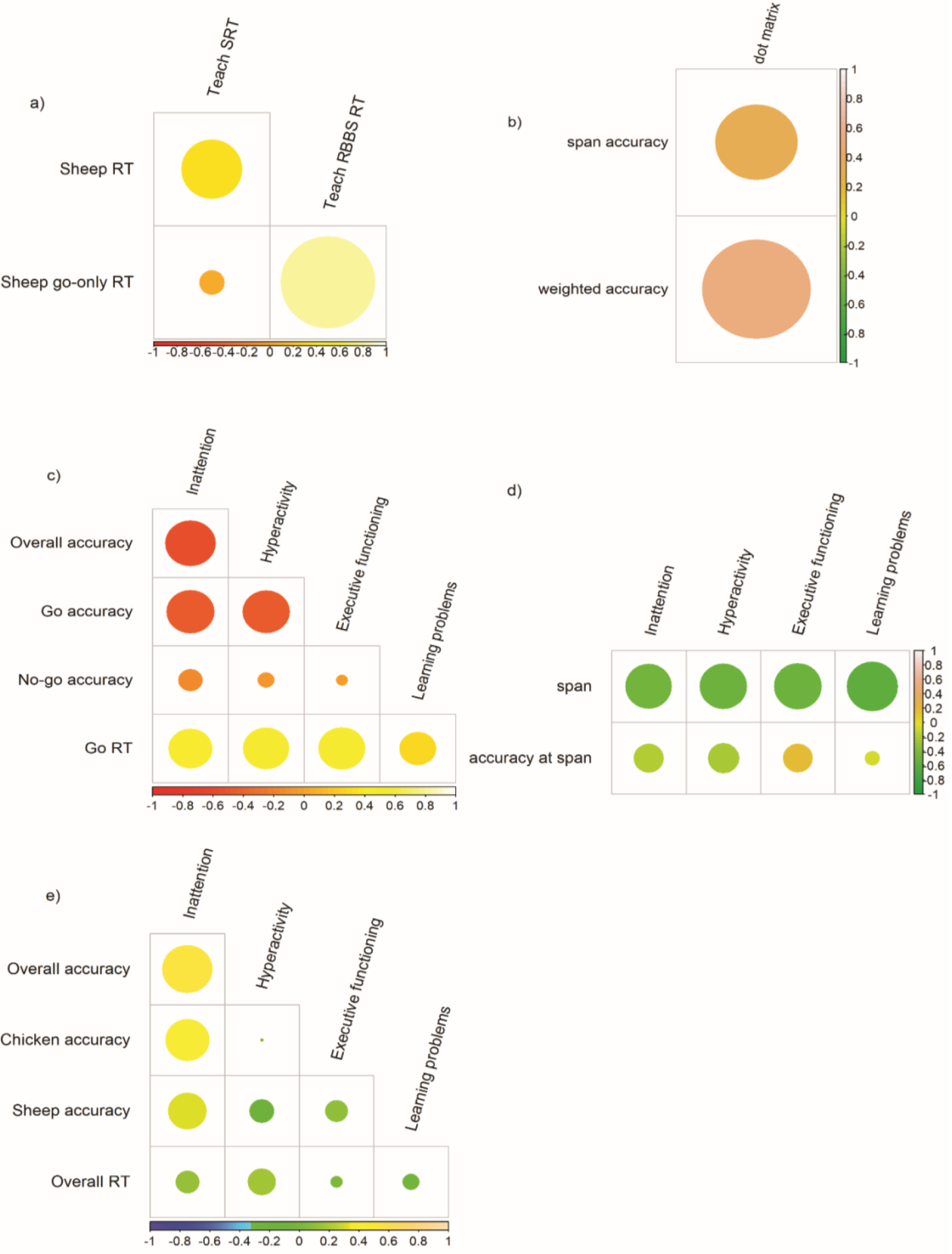
Correlation between FarmApp performance, neuropsychology tests, and ADHD traits

### 2b. Validation against behavioural questionnaires

Here, we explored whether FarmApp game performance was associated with everyday behavioural characteristics (ADHD questionnaire scores), within the whole sample (BINGO and CALM). We focussed on the performance in Child mode across all three games (Sheep, Chicken, and Memory), applying the same inclusion criteria and performance metrics as previously described. Correlation matrices are illustrated in Figure 2 (panels c, d, e). After covarying for age and Vineland ABC, the following associations were observed. For the Sheep game (N=45), go-only accuracy negatively correlated with inattention (*r*_*s*_=.425, p<.02), and executive functions (*r*_*s*_=-.462, p<.008); i.e. children who were better at responding to targets in the Sheep game exhibited fewer parent-reported difficulties with inattention and executive functions. In the Chicken game (N=33), span and highest level reached negatively correlated with inattention (*r*_*s*_=-.377, p<.04) and hyperactivity (*r*_*s*_=-.481, p<.007) after covarying for age and Vineland, i.e. better performance at the Chicken game was correlated with fewer parent-reported difficulties with inattention and hyperactivity. For the Memory game (N=14), overall accuracy positively correlated with inattention (*r*_*s*_=753, p<.003) after covarying for age and Vineland; i.e. participants most likely to distinguish “previously seen” from “new” items were reported to have more inattention symptoms by parents.

#### FarmApp and neuropsychological batteries

2a) Correlation matrix showing the relationship between performance at the Sheep game (go-only and mixed RT) and Teach2 subtests (RBBS and SRT). The colour scale goes from red, negative correlation, towards positive, or yellow, Spearman Rho values. The size of the circles is the magnitude of the effect, or p-values. 2b) Correlation matrix between the performance at the chicken game (span and weighted accuracy) and AWMA Dot matrix subtest. The correlation matrices here illustrated have not been controlled for age and general ability.

#### FarmApp and behaviour

2c) Correlation matrix between performance at the Sheep game (accuracy and RT) and ADHD traits. The colour scale goes from dark red, negative correlation, towards positive, or yellow, Spearman Rho values. The size of the circles is the magnitude of the effect. 2d) Correlation matrix between performance at the Chicken game and ADHD traits. 2e) Correlation matrix between performance at the Memory game and ADHD traits.

### 3. Change over time

The third part of the study explored change in attention task performance over time (Child mode, Sheep game) over a two-week period of time, for the combined CALM and BINGO samples (N=48). We examined learning curves, and focussed our analysis on the no-go stimulus duration, as an adaptive test parameter reflecting improvement in the ability to respond to targets and inhibit the response to distractors (see supplementary material for participant-level raw data). After excluding participants (n=12) whose performance resulted significant at the randomness test (*runstest* in Matlab), the analysis was performed by fitting linear mixed effects models (LMMs) with *lmer* toolbox in R. Two linear mixed effects models or LMMs were fitted to the data, where the first model included general ability (Vineland ABC) as an interaction factor, and the second set of models included inattention, hyperactivity, and EF (ADHD questionnaire scores). Summaries of model fitting are presented within supplementary material.

After centering our variables of interest (around the mean), we defined a linear model with random slopes and intercepts. In this case, the number of blocks, age, and interaction between number of blocks and level of general ability were fitted as fixed factors, allowing the slopes to vary across subjects and age. A total of 48 participants were included in this analysis. There was a significant effect of the number of blocks played (β= −9.98, CI= −18.90 - −1.5), meaning the more children played the lower the stimulus duration dropped. In addition, there was a significant interaction between general ability and the number of blocks played (Block N x Vineland, β=14.38, CI= 3.80-24.96). The results of this LMM, including general ability (Vineland composite scores) as the interaction factor, suggest that children did improve in their ability to inhibit the response to the distractor stimuli, and that this improvement was greatest for children of lower general ability, who started with a longer no-go stimulus duration. Amongst more able children, the test parameters did not enable them to improve over time, and we observed a tendency in their performance to deteriorate rather than to improve (Figure 3a).

**Figure 3.**
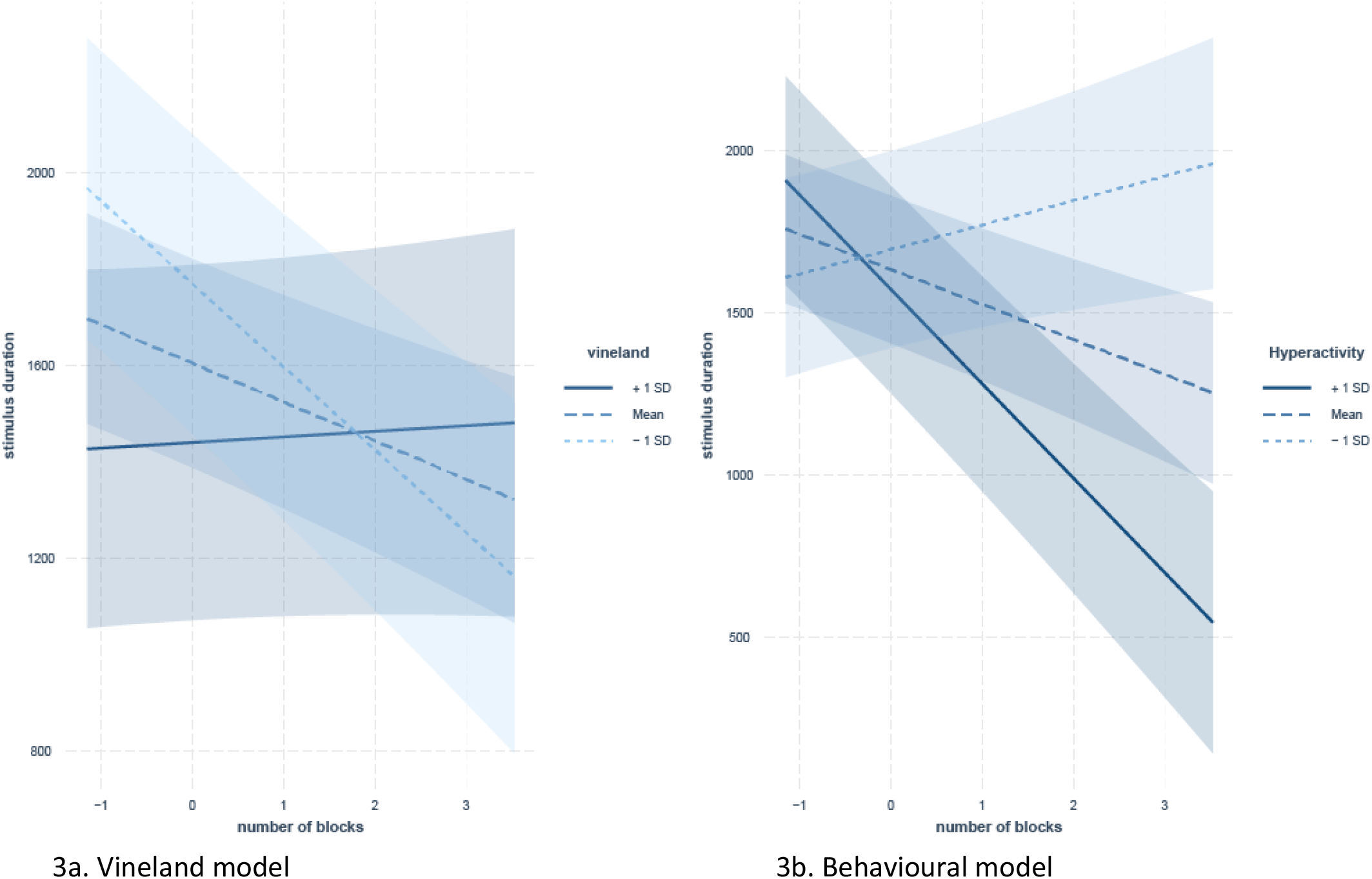
Mixed effects linear models including general ability (Vineland) and hyperactivity as interaction factors. 3a) Relationship between general ability (Vineland ABC) and change in Sheep game no-go stimulus duration across number of blocks. 3b) Relationship between hyperactivity and change in Sheep game performance (no-go stimulus duration) across number of blocks.

The second set of models included Conners subscale’s scores of Hyperactivity, Inattention and Executive Functions as interaction terms, with block number and age as fixed factors, plus subjects as random factor. Overall, 35 participants were included in this analysis. The results showed significant main effect of number of blocks played on the stimulus duration estimate (β= −118.95, CI=-162.01 – - 75.88), as well as a significant interaction between hyperactivity and the number of blocks played on the no-go stimulus duration prediction (Block N x Hyperactivity, β=-108.60, CI= −182.24 – −34.96). Following the behavioural LMM, we were interested in the predictive power of each of the ADHD traits in the model fit. For this reason, we fitted three separate LMMs (hyperactivity, inattention, and EF) and compared the models with ANOVA and information criteria metrics (AIC, BIC). While in each of the single LMMs there was a significant interaction effect with the number of blocks played, the best explaining model of the data, based on the ANOVA and the AIC and BIC criteria, included hyperactivity as the interaction factor (AIC < 2). These results show how higher hyperactivity (i.e. + 1 SD above the group mean) was associated with steeper learning curves, where children improved more at the Sheep game, the more they played. Interaction effects of each of the models are shown in supplementary material.

### 4. Effects of genetic diagnosis on FarmApp performance and change over time

Lastly, we addressed the hypothesis that FarmApp performance is influenced by genetic diagnosis within the BINGO sample of individuals with ID of known genetic origin. The BINGO sample (N=67) was divided into two groups, based on the functional network of genetic diagnosis: a chromatin-related group, and synaptic-related group. Table 3 presents descriptive data for the two groups, and comparison of their performance on FarmApp games (Child mode, collapsed over time).

**Table 3.** Summary table of the comparison between gene functional network groups.

|  | Participants |  | Chromatin | Synaptic | Test statistic |
| --- | --- | --- | --- | --- | --- |
| Number | 67 |  | 27 | 30 |  |
| Age | Mean (range, SD) | 67 | 149<br>(69.0-306, 59.9) | 160<br>(1-315, 77.1) |  |
| Gender | Female:Male | 41:26 | 11:16 | 30:10 |  |
| Vineland ABC | Mean (range, SD) | 67 | 64.2<br>(33.0- 96.0, 12.7) | 49.4<br>(20.0-69.0,12.9) | **p<.001** |
| Sheep<br>(Child mode) | Overall accuracy | 42 | 0.758<br>(0.580- 0.870, 0.0852) | 0.658<br>(0.440- 0.860, 0.122) | **F(1,38)=4.35,**<br>**p=.031** |
|  | Go accuracy | 42 | 0.758<br>(0.580-0.870, 0.0852) | 0.658<br>(00.440-0.860, 0.122) | **F(1,38)=2.87,**<br>**p=.036** |
|  | No-Go accuracy | 42 | 0.801<br>(0.365-1.00, 0.159) | 0.744<br>(0.314-1.00, 0.177) | F(1, 38)=2.29 |
|  | Go RT | 42 | 793<br>(573-1460, 201) | 1200<br>(696-2040, 420) | **F(1,38)=10.73,**<br>**p<.01** |
| Chicken<br>(Child mode) | Span | 42 | 4.26<br>(2.00-8.00, 1.45) | 3.09<br>(1.00-7.00, 1.57) | F(1, 37)=1.88 |
|  | Span accuracy | 42 | 0.239<br>(0.0600-0.420, 0.0993) | 0.179<br>(0.0200- 0.580,0.126) | F(1, 37)=0.92 |
| Memory<br>(Child mode) | Total accuracy | 17 | 0.734<br>(0.490-0.980, 0.183) | 0.632<br>(0.480-1.00, 0.175) | F(1,13)=1.046 |
|  | Total RT | 17 | 1950<br>(922-4700, 1060) | 2040<br>(1250-3720, 870) | F(1, 13)=1.88 |

Analyses of FarmApp variables covarying for age and Vineland ABC. In bold the significant differences, Bonferroni corrected.

First, we compared Child mode performance on all three games between the two groups, covarying for age and Vineland ABC. The results for the Sheep game show that the chromatin group were significantly more accurate (overall accuracy) and faster to respond to go-only trials (Go RT). However, there were no differences between groups in Chicken game or Memory game performance (Table 3).

Next, in order to examine whether the genetic group had an effect on learning slopes in the Sheep game, we fitted a LMM including age, Vineland ABC, and an interaction between block number and genetic group, with subject and age as random factors (model fit results are shown in supplementary material). Overall 35 participants were included in this analysis. The results showed a significant effect of the block number i.e. as children played the game more, they improved, hence the no-go stimulus duration dropped (Block number, β= −113.99, CI= −176.28 – −51.69). In addition, there was a significant effect of genetic group on the relationship between number of blocks played and no-go stimulus duration (Block N x Genetic group, β=-341.69, CI= −458.51 – −224.87). Figure 4 illustrates the group-wise changes in performance over time - the synaptic group showed a steeper rate of improvement over time, whereas the chromatin group, while also improving, showed a flatter slope.

**Figure 4.**
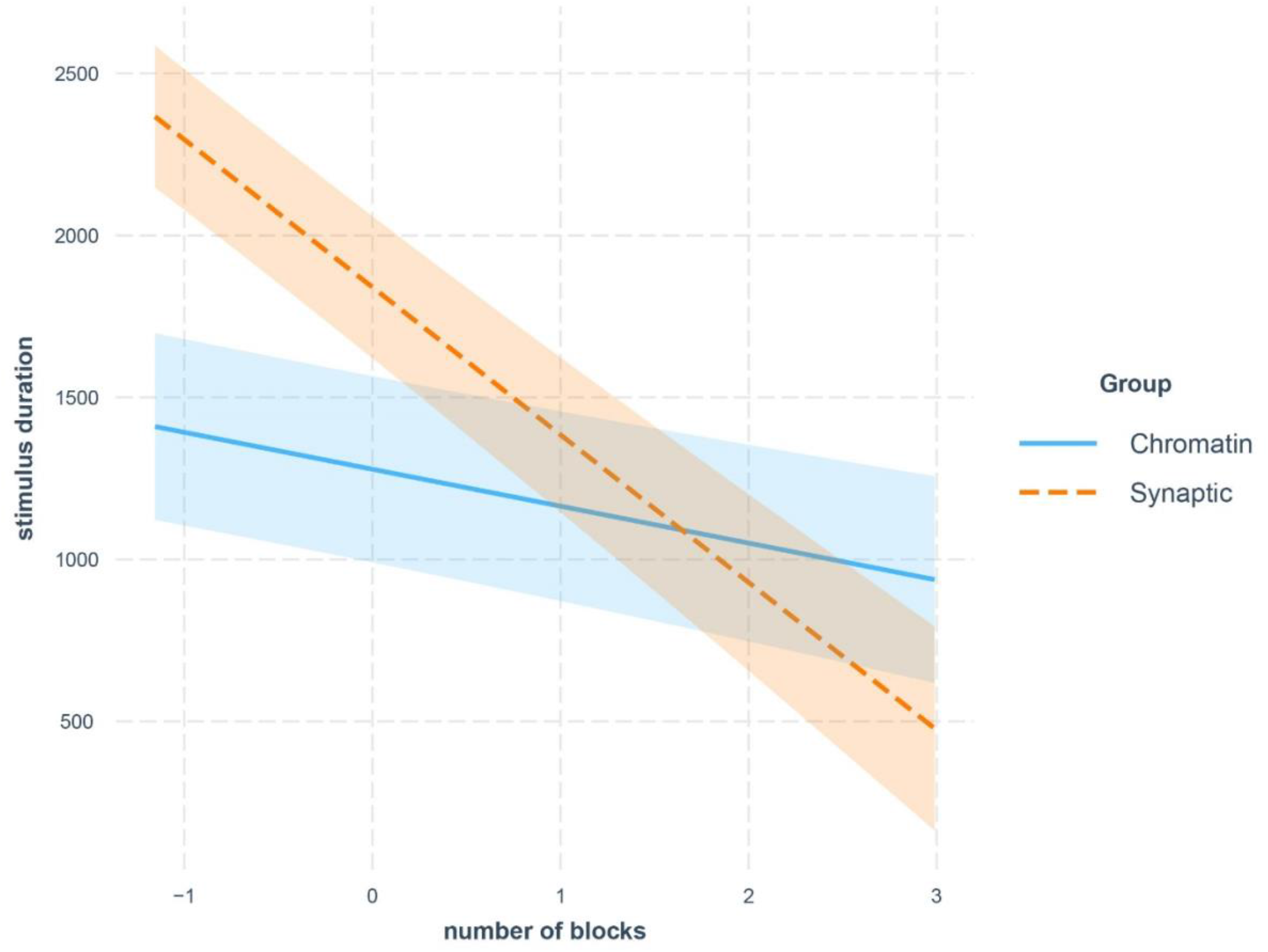
Interaction effect of functional genetic networks group in the model fit. This figure shows the interaction effect (with 80% CI) of group in the model fit. The Synaptic group had a higher intercept, or started slower, and improved more in the game, when compared to the chromatin-related group (blue).

## Discussion

The current study proposes an alternative to traditional cognitive assessments for individuals with ID. We designed a new touchscreen-based app as an accessible assessment method that measures stimulus response, speed, selective attention, short-term and long-term memory capacity. A key element is the encouragement of repeated testing and analysis of performance changes over time. The overarching goal of the current research was to examine FarmApp’s potential to measure cognitive abilities across a wide age range and spectrum of neurodevelopmental disorder severity. We address four research objectives: feasibility, validity, analysis of change in performance over time, and the utility of FarmApp for linking causal factors such as genetic diagnosis to cognitive mechanisms.

### Feasibility

We observed that more than half of recruited participants were able to provide analysable data in RA mode, but this proportion increased in Child mode, when children could play at their own time and pace. Many children in the BINGO sample struggled to complete a standardised cognitive assessment (Leiter or WASI-II), but found FarmApp intuitive and enjoyable by comparison. This suggests that to capture cognitive competence in individuals with ID, traditional one-off neuropsychological testing is suboptimal. Remote assessment via FarmApp can yield multiple samples of performance and add opportunities to observe fluctuations in cognitive performance over timescales of hours, days and weeks. However, younger children and participants with more severe global impairments were less likely to provide sufficient data for analysis of tasks. This was especially relevant for the recognition memory task which requires a minimum threshold of play and exposure to stimuli before becoming available on the App. Future improvements to the App will seek to address these limitations and broaden accessibility further, whilst retaining adaptive elements of the design to maintain motivation and avoid ceiling effects for more able participants.

### Validity

Sheep and chicken RA mode performance (accuracy and RT) positively correlated with equivalent measures of attentional performance and spatial working memory from the Tea-Ch2 and AWMA batteries. This suggests that the App platform is sensitive to cognitive processes tapped by standardised tests appropriate for higher ability children. We acknowledge that only a few metrics from the FarmApp games, such as RT in the Sheep game and span in the Chicken game, were directly comparable to raw score metrics from the Tea-Ch2 and AWMA batteries. A limitation that is difficult to overcome is that this correlational analysis of validity was only possible in the small group of higher ability participants from the CALM cohort in whom the standardised tests were accessible. An alternative might be to align the standardisation of measures of accuracy in all three games (sheep, chicken, memory) with parallel measures from other batteries, and pursue larger scale testing and normative data collection across different ages and cognitive abilities.

Moreover, in the combined BINGO and CALM studies, FarmApp performance related to parental reports of everyday difficulties assessed via a standardised ADHD questionnaire. Specifically, both sheep and chicken performance metrics correlated in the expected direction with symptoms of inattention, hyperactivity and executive dysfunction. These results suggest that FarmApp taps cognitive processes contributing to everyday behaviour, and can be a useful tool for understanding mechanisms contributing to behavioural difficulties in young people with ID. A surprising result was the association between better memory performance and higher inattention symptom scores – the most likely explanation is that individuals with inattention were less likely to progress through adaptive phases of the games, meaning that stimulus presentation rates remained lower and stimuli were presented to these participants for longer durations, and were thus more likely to be recognised. Although this is a limitation of the study design, the observation could have simple translational implications, highlighting the need to present to-be-learned information at a slower rate and for longer time for individuals with attention difficulties.

### Change over time

In order to assess whether FarmApp can track dynamic changes in game performance over time, we focussed our analysis on the Sheep game, because of the large amount of data provided by participants. By fitting LMMs we explored the relationships between time and performance, and the potential contributions (or interaction) of participants’ age and general ability to change in performance. When allowing our models to vary for age and across subjects (random effects), we found a significant interaction of general ability (Vineland) on the relationship between number of blocks and the improvement at inhibiting the response to the no-go stimuli (indexed by stimulus duration). Although this shows that FarmApp was an effective measure of improvement in participants with lower cognitive ability, children with higher cognitive ability quickly adapted their performance, with smaller range available to demonstrate longer-term improvement. This is an important finding to be integrated in the next version of FarmApp: the training and warm-up phases might be used to establish individual threshold and cut-off of improvement, by increasing the range of stimulus presentation rates in the Sheep game, and the number of hutches in the Chicken game. Furthermore, we explored whether behavioural traits would have an effect on change in performance, or predictive value in our model estimates. We found that hyperactivity was associated with progressively better inhibitory ability, or lower stimulus duration over blocks. Albeit counterintuitive, this outcome highlights how children who might have higher rates of behavioural problems, are still able to improve at FarmApp games. In other words, FarmApp could be a sensitive tool for measuring change in cognitive performance in individuals with lower cognitive abilities and higher levels of behavioural difficulties. An alternative interpretation is that for those individuals with lower adaptive ability, greater ADHD traits and poorer baseline performance, there was more scope for improvement at the game over time. Specifically, longer term testing enabled these participants to demonstrate their “best possible” performance. In contrast, individuals with higher adaptive ability, lower ADHD traits, and better baseline performance may have been at or close to ceiling level at the outset, and therefore improved only marginally over the two-week period. One way to explore which of the two alternative interpretations is more valid would be to carry out a separate study comparing change in performance over time in children diagnosed with ADHD and age-matched controls. Future studies could explore how fluctuations (beyond improvement with practice) in specific aspects of FarmApp performance within an individual may mirror their variation in engagement with education, social interactions, or emotional tone. Potentially this could highlight opportunities for targeted interventions to improve extrinsic modifiable factors such as sleep and diet modification, or neurochemical modulation targeted at each individual’s most vulnerable cognitive skills.

### Causal factors and cognitive mechanisms

Finally, to assess the utility of the method for linking causal factors and cognitive mechanisms we compared FarmApp task performance between individuals with ID due to different genetic variants, grouped according to gene function (16). We found that, after controlling for age and general ability, the chromatin-related group performed significantly better than the synaptic-related group on the selective attention task, but groups did not differ in spatial short-term memory or longer term recognition. These findings are in line with current models of synaptic regulatory mechanisms and genetic disorders and their effects on attentional control (29, 30). For instance, poorer performance in tasks requiring inhibitory control have been reported in Fragile X Syndrome (FXS), in children and adults, supported both in cross-sectional and longitudinal studies (15, 31-33). Our speculation is that synaptic disorders have a direct and continuous impact on cognitive processes requiring fast information processing and adaptation of responses over short (millisecond to seconds) timescales. Chromatin-related disorders are more likely to mediate their impact on cognition and learning over longer timescales via altered gene transcription and slower adaptations to neuronal biology and neural network function, of maximal importance during critical periods of brain development. Our results support the general hypothesis that multiple cognitive processes contribute to learning in neurodevelopmental disorders, and that genetic disorders (and functional network groups) can have a disproportionate, if not selective, impact on specific cognitive processes.

In addition, whilst the chromatin-related group’s performance stayed relatively stable, the synaptic-related group improved over blocks. Similar to the whole sample findings on change over time, this may reflect differential capacity for improvement on this task for individuals with synaptic-related disorders and more severe ID, who have more difficulties performing the task at baseline. Alternatively, different functional networks might influence different physiological mechanisms, contributing to the acquisition of skills versus potential competence level versus skill maintenance. Our results could suggest that individuals with synaptic-related disorders have equivalent potential competence level to the chromatin-related disorders group, but require more opportunity to acquire competence and reach their maximal ability on this task. A similar trend has been observed in another synaptic related syndrome, where in a three-year perspective study FXS boys improved in their visual attentional cognitive control over time (15). In summary, our last analysis showed how FarmApp can be applied to investigate genetic diagnosis-influenced change in cognitive performance.

## Limitations and future directions

The results described here do come with a list of limitations. First, the power of our results is limited by the relatively small sample size, especially in the genetic analysis. Although the current findings represent an important step forward into testing alternative methods for assessing cognitive competence in ID, further research involving multicentric and international recruitment and data collection would allow to capitalise on FarmApp’s potential as a valid and effective computerised tool.

Furthermore, the utility of assessing performance over time is highly dependent on the statistical methods employed to measure dynamic change in performance. Keeping this in mind, we have applied linear mixed models that take into account between and within subjects’ variability (28). The amount of time allowed for longitudinal data collection (two weeks of free play) might not have been enough to observe change in performance for all participants. One way to overcome this issue would be to systematically monitor whether changing the duration of the longitudinal data collection (e.g. two-weeks, a month, or longer) would have an impact on fluctuations in performance. There was also a high degree of variability in amount of data acquired for each participant during this time, a limitation inherent to participant-led assessment. This variability could be systematically investigated with more participants and more data, allowing thus to identify potential cognitive and behavioural contributing factors.

Finally, when measuring change in performance over time we have consistently focused on inhibitory control during the visual attention task (no-go stimulus duration). This metric was chosen as the most sensitive measure of improvement, as a trade-off between accuracy at the go-only trials and preparedness to inhibit distractors. Future development of FarmApp is needed to facilitate dynamic analysis of short term and longer-term memory tasks, and to be able to expand the utility of the method in both research and clinical settings.

## Conclusion

The present study offers an example of an alternative cognitive assessment platform, intuitive, easy to use, and most of all, feasible for individuals who have difficulties attending to and understanding task instructions. Moreover, FarmApp has the additional benefit of offering longitudinal monitoring of cognitive performance over time. Current findings represent a promising avenue for the use of touchscreen tools and remote long-term assessment, to systematically investigate dynamic aspects of development. Future studies will provide new insights into the relationships between cognitive processing and behavioural variation over different timescales, and the influence of specific genetic diagnosis on emergent cognitive differences.

## Data Availability

The data that support the findings of this study are available from the corresponding authors upon reasonable request.

## Declarations

## Acknowledgments

We sincerely thank the study participants, their families and carers for the contribution to this study. The research team acknowledges the support of referring clinicians and the National Institute for Health Research, through the Comprehensive Clinical Research Network and Rare Genetic Disease Research Consortium. We are also grateful to Ounce technologies (https://ouncetech.co.uk/projects/) for supporting the design and implementation of FarmApp.

## Ethics approval and consent to participate

Parent or carer provided written informed consent on behalf of participants under the age of 16. Ethical approval for the CALM study was granted by the National Health Service (NHS) Health Research Authority NRES Committee East of England, REC approval reference 13/EE/0157, IRAS 127675. Ethical approval for the BINGO study was granted by the NHS Health Research Authority NRES Committee East of England, REC approval; reference 11/EE/0330, IRAS 83633.

## Author’s contribution

DA, KB and GS conceived the study. ENC and SOB acquired the data. DB and ENC analysed and interpreted the data. JM provided the figures of the FarmApp game structure. DB, KB and ENC drafted the paper and all the other authors substantially revised it. All authors read and approved the final manuscript.

## Open Practices Statement

This article is distributed under the terms of the Creative Commons Attribution 4.0 International License (http://creativecommons.org/licenses/by/4.0/), which permits unrestricted use, distribution, and reproduction in any medium, provided you give appropriate credit to the original author(s) and the source, provide a link to the Creative Commons license, and indicate if changes were made.

