## supplementary material for "FarmApp: a new cognitive assessment method for young people with intellectual disability"

### Game structure

Below is the detailed description of the FarmApp game structure. In order of game administration, sheep, chicken, and memory game.

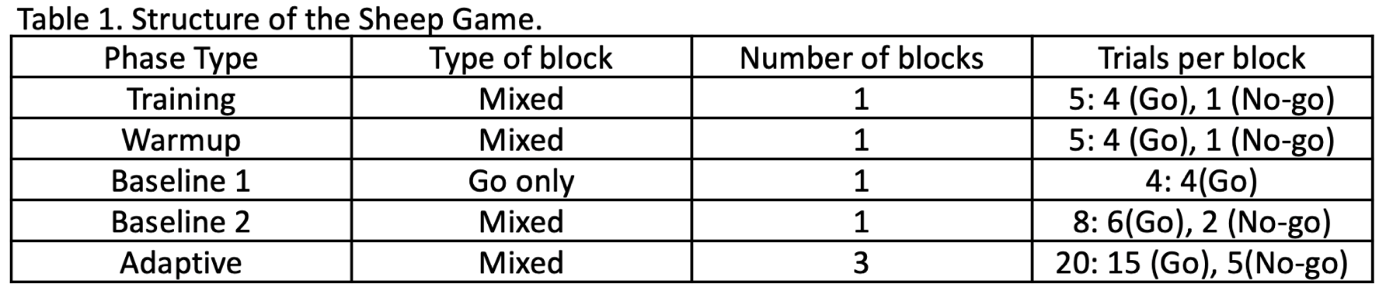
**Sheep game:** this game is a Go/No-Go task comprised of five phases: training, warmup, baseline1, baseline2, and adaptive. The game is introduced by an audio trailer and participants were told that the naughty pigs had let the farmer’s sheep out of their field and that they must help the researcher collect the sheep. They were instructed to touch the sheep, not the pigs. This was demonstrated to the child in the training phase. The child then completed the other four phases of the task (Table 1.), independently with encouragement from the researcher.

Table 1. Structure of the Sheep game.

Each trial begins with the presentation of a fixation screen. In Go-only trials (Figure 1B) a sheep stimulus, in different costumes across trials, was presented in the bottom left-hand corner of the screen, whereas in No-go trials (Figure 1C) a pig stimulus was presented. In the adaptive phase, the response window duration varied with the child’s ability. A correct response in a Go-only occurred when a child touched the sheep stimulus, generating a ‘Baa’ sound and the sheep stimulus moving to the top right-hand corner of the screen. In contrast, a correct response in a No-go trial occurred when the child did not touch the pig stimulus (inhibitory response) and resulted in an ‘Oink’ sound. Additionally, either trial could finish with an incorrect or no response, generating no feedback. The progression of the game was shown to the child after each block by audio and visual feedback of a dog barking and moving along the bottom of the farm-scape towards a kennel (Figure 1D). Once all the blocks were complete, the end of game screen was presented (Figure 1E).

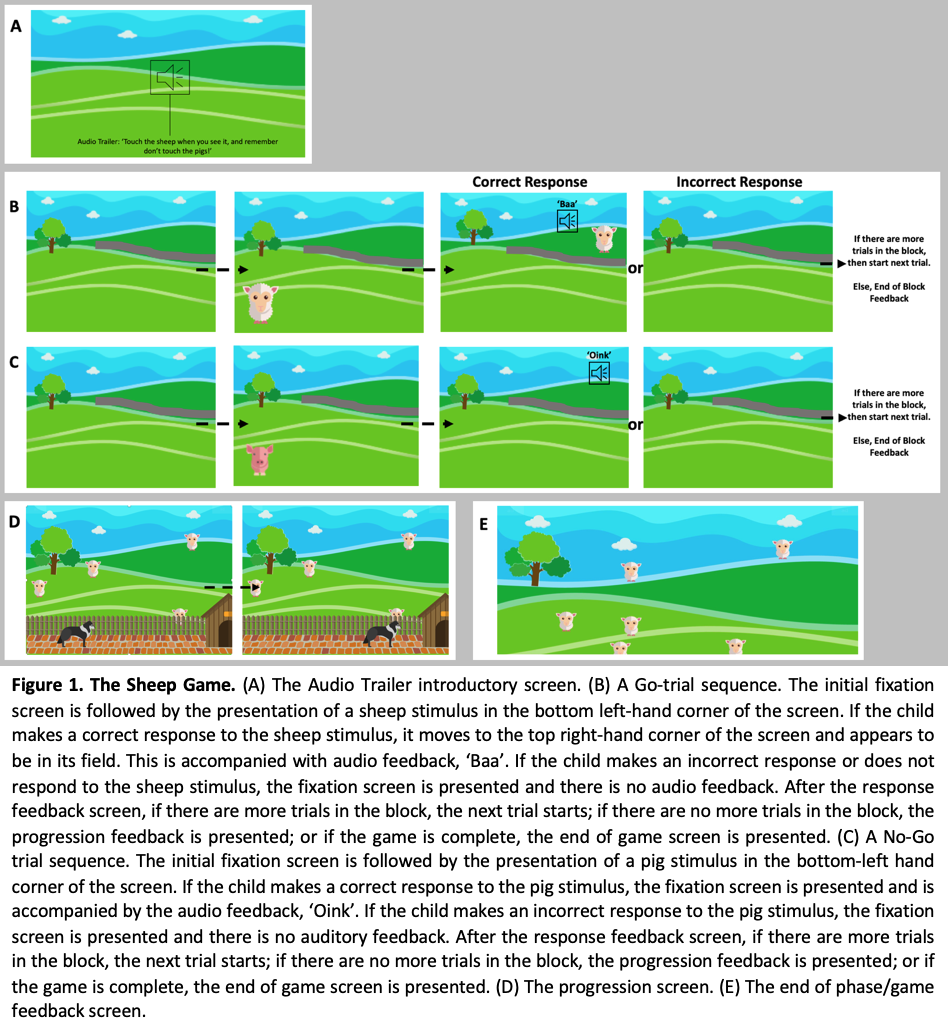
Figure 1. The Sheep Game. A) The Audio trailer introductory scene. B) Go-only trial sequence. The fixation green screen is followed by the presentation of a sheep stimulus (second frame from the left in the B row) in the bottom left-hand corner of the screen. If the response is correct, by tapping on the sheep, an audio feedback (“Baa”) is produced. In case of missed or incorrect response the fixation screen is presented without audio feedback. At the end of the block or the game, the progression feedback (the dog barking). C) No-go trial sequence. Same as for the go-only trials, in the no-go trials a pig is presented in the bottom left corner of the screen. If the response is correct, the fixation green screen is accompanied by the audio feedback sound (“Oink”). In case of missed or incorrect response, the fixation screen is presented with no audio feedback. D) The progression screen. E) The end of phase (go-only, mixed) and end of game screen.

**Chicken game:** Same as for the Sheep game, the game has four phases: training, warmup, baseline, and adaptive. An audio trailer introduces the Chicken game (Corsi-block test), where participants are told that the chickens on the farm were laying eggs and they must help the researcher collect them for the farmer. Participants are instructed to carefully watch the order the chickens emerge from their hutches and to collect the eggs they must touch the chickens in the same order they remember seeing them. This was demonstrated in the training phase. The child then completed the other three phases of the task: a warm-up, baseline and adaptive phase, independently with encouragement from the
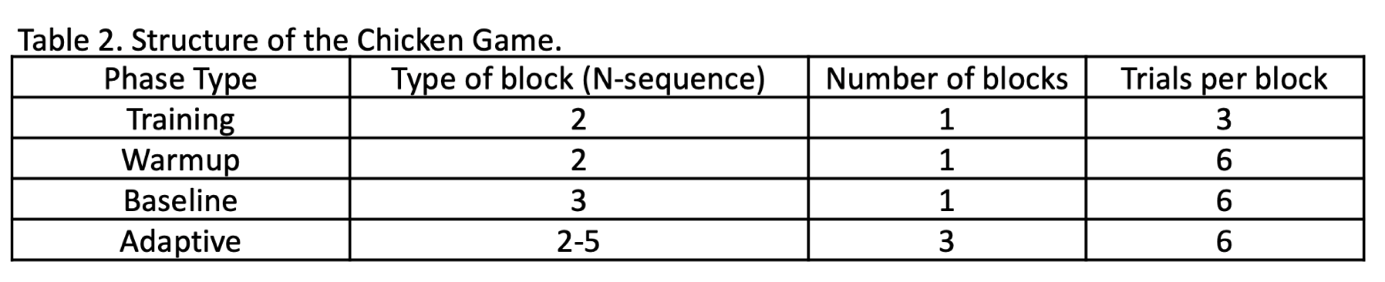
researcher (Table 2).

Table 2. Structure of the Chicken game.

Each trial starts with the fixation screen containing eight chicken coups (Figure 2B). This is followed by a sequence of chicken stimuli, ranging from 2-5 chickens, in varying costumes ‘popping-out’ of hutches. Each stimulus is accompanied by the audio ‘chicken cluck’, when they appeared. In the adaptive phase, the length of the sequence or chicken span is based on participant’s ability. An audio instruction ‘Go’ indicated that the child should respond after the last chicken of the sequence is presented. A trial could end in four ways: 1) A correct response, where all the coups had to be tapped in the correct order. In this case, a visual and audio feedback of a chicken laying an egg occurs when the last hutch in the sequence is tapped. 2) Incorrect response where ‘Whoops, let’s try again’ as audio feedback is presented, as soon as an incorrect tap happens. 3) A trial could also end if a response did not occur in the initial no response timeout duration or; 4) if not all the responses occur within the no response timeout. The progression of the game is presented to the child by audio and visual feedback of a rolling sound and an egg moving towards a basket (Figure 2C). Once all the blocks are complete, the end of game screen was presented (Figure 2D).

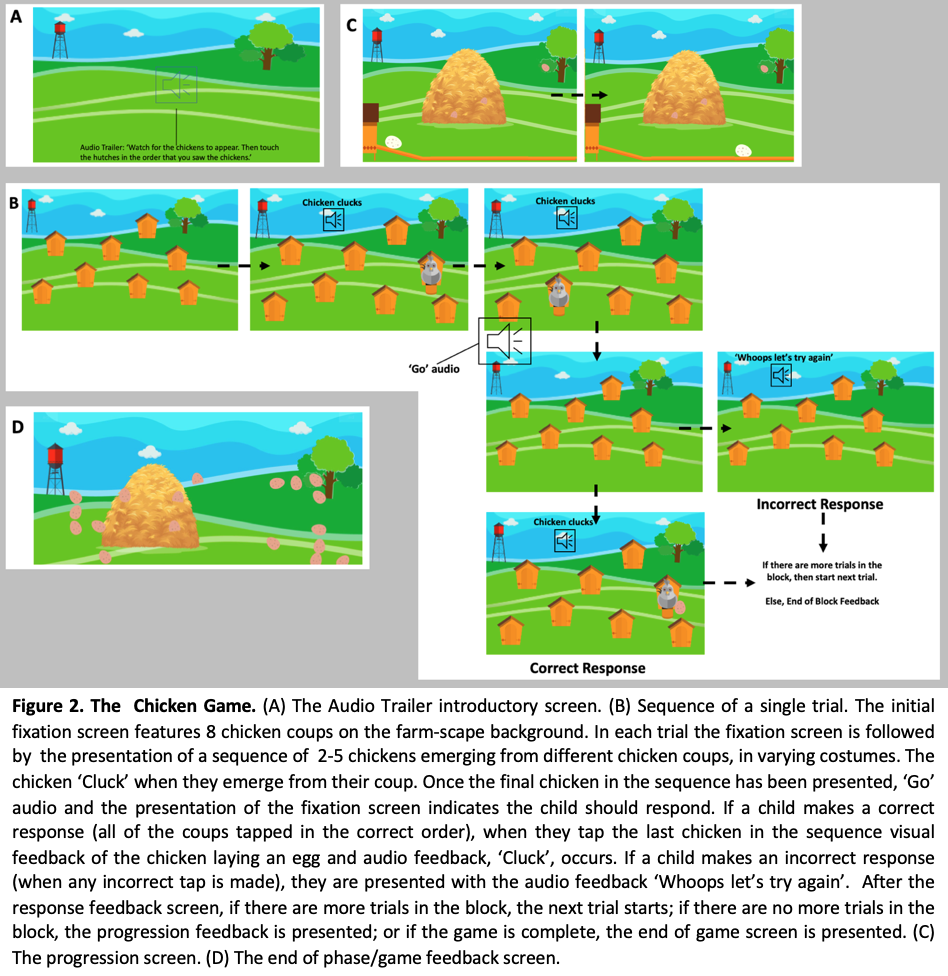

Figure 2. The Chicken game. A) The introductory scene with the audio trailer. B) Single trial sequence. The first fixation screen features eight chicken in hutches in the farm-scape background. In each of the trials this is followed by the presentation of the chicken in various costumes, the number of chicken varies between 2-5 in RA mode, and in child mode may go up to eight chicken. After the last chicken presentation of the sequence (trial) an audio prompt “Go!” indicates to the participant to start tapping the coups in the same exact order as the stimulus sequence presentation. In case of correct response, or all the hutches tapped in the correct order, an audio-visual feedback is presented with the image of the chicken laying an egg and a “Cluck” sound (C). In the case of incorrect tap, a different audio feedback is heard (“Ops! Let’s try this again!”). C) The progression screen. D) The end of a phase or game.

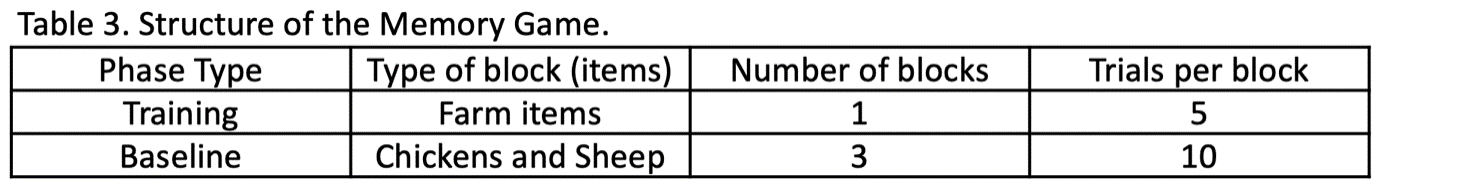
**Memory game:** The final game is longer-term recognition memory test, composed of two phases: training and baseline (Table 3). Participants are told, that since they have played the previous sheep and chicken game, now they need to try to remember what they have seen. When presented with pairs of either chicken or sheep, they need to choose which one they have seen before, by tapping on the character that looks familiar (Figure 3).

Table 3. Memory game structure

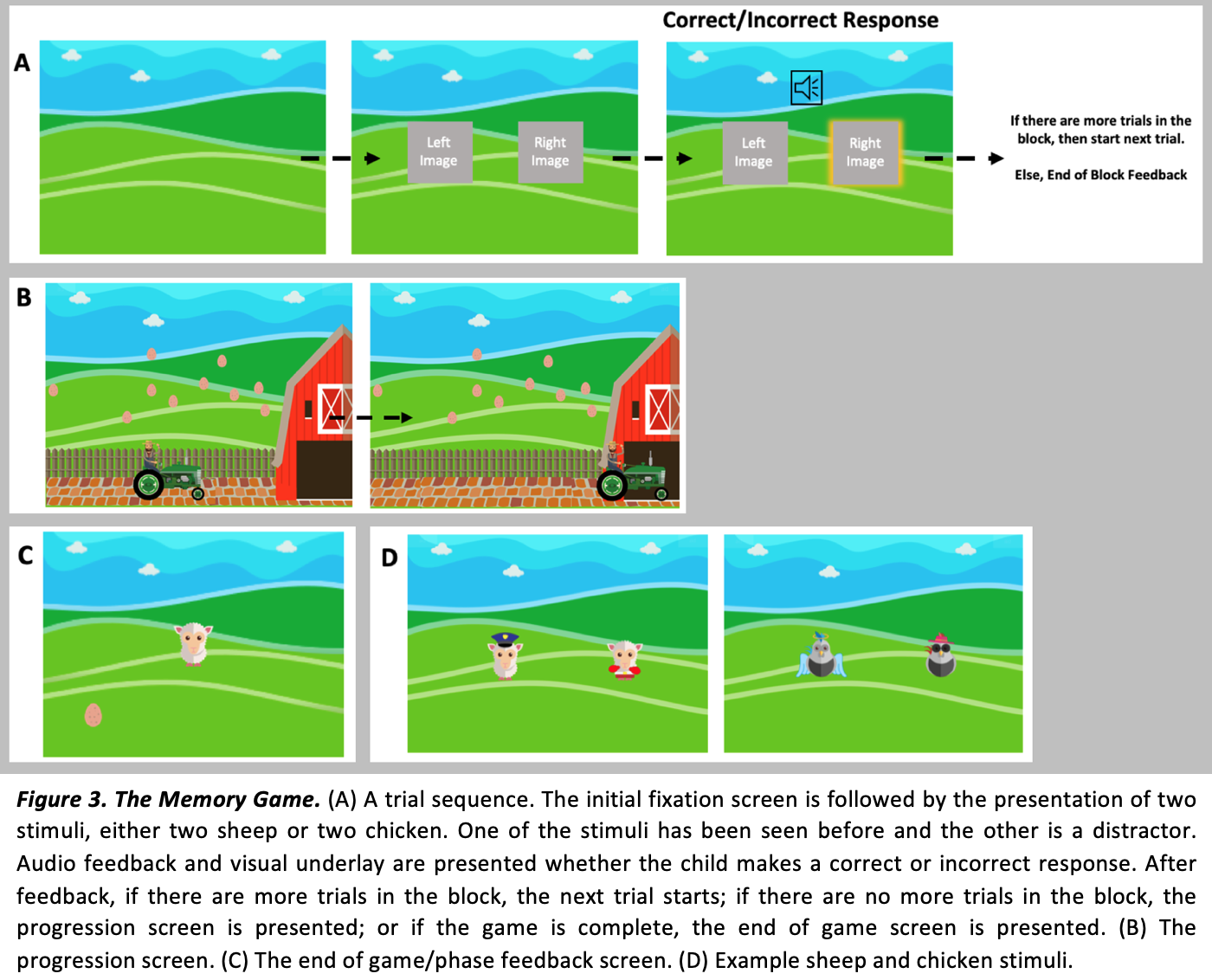
 In the training phase, to avoid stimuli familiarisation, items from the farm-scape were used to prompt participants’ items recall (e.g. dog kernel). Following the successful training phase, participants completed the baseline phase of the task independently.

Figure 3. The memory game. A) A trial sequence; the initial fixation screen is followed by the presentation of a pair of stimuli, with either two sheep or chicken. D) One of these has been presented in the two previous games and the other is a distractor. Audio feedback and visual underlay are always presented, no matter whether the response is correct or incorrect. B) At the end of the block the progression screen is presented. C) End of the game screen.

### Inclusion criteria for RA and Child mode games

**Inclusion criteria for each game in RA and Child mode:** For both modes analysed in the attached manuscript, RA and Child mode, the inclusion criteria were: for the sheep game minimum of five no-go trials, equal to at least one block completed; in the chicken game participants who completed at least one level of hutches (or block), were included in the analysis; for the memory game participants who had one completed run were included in the analysis.

**Inclusion criteria for change over time study:** For the two studies we looked the at change in performance over time in the Sheep game, by applying linear mixed modelling (LMMs) to the no-go stimulus duration as a measure of performance. Here, since the data was in the long format, we performed the *runtest* in Matlab, for both no-go stimulus duration and go-only RT. *Runtest* returns a test decision for the null hypothesis that the values in the data come in random order. The test is based on the number of runs of consecutive values that are either above or below the mean. The participants whose performance at both was no random, were included in the analysis.

### Study 3: Change over time

Illustrated in Figure 4 is the individual fitted data, or learning curves of individual subjects (stimulus duration) plotted against time or number of blocks.

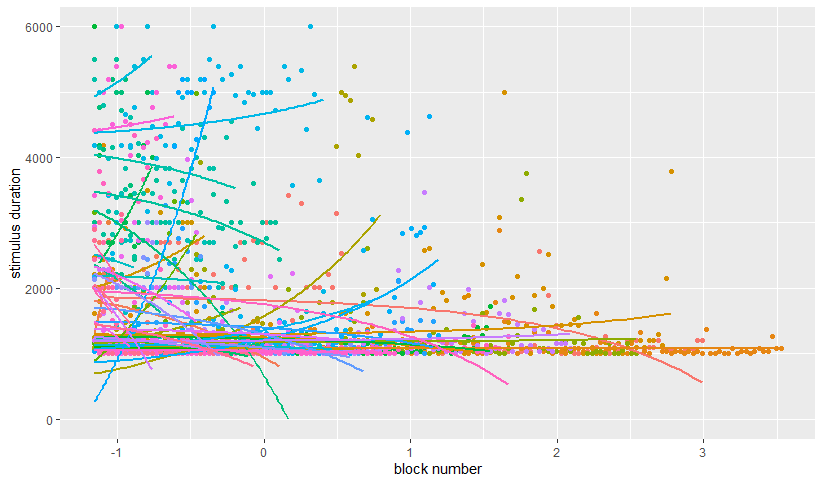

Figure 4. Individual fitted data, based on the number of blocks played (centred around the mean), each colour pictures individual subjects. This plot includes all the subjects (CALM + BINGO) included in the analyses illustrated below.

#### Vineland model

In Table 4, summary of the first LMM having as predictors: number of blocks, general ability measure (Vineland), age, and the interaction between measure of general ability (Vineland) and the number of blocks played.

|  | **Stimulus duration** | | |
| --- | --- | --- | --- |
| *Predictors* | *Estimates* | *CI* | *p* |
| (Intercept) | 1655.63 | 1434.18 – 1877.09 | **<0.001** |
| Block number | -9.98 | -18.90 – -1.05 | **0.029** |
| Vineland | -212.00 | -479.67 – 55.67 | 0.132 |
| Age | -214.46 | -603.98 – 175.06 | 0.289 |
| Block nr:Vineland | 14.38 | 3.80 – 24.96 | **0.008** |
| **Random Effects** | | | |
| σ^2^ | 407041.27 | | |
| τ_00_ _subject_ | 236656.49 | | |
| τ_11_ _subject.age_months_ | 755260.02 | | |
| ρ_01_ _subject_ | -0.01 | | |
| ICC | 0.71 | | |
| N _subject_ | 48 | | |
| Observations | 2173 | | |
| Marginal R^2^ / Conditional R^2^ | 0.026 / 0.717 | | |

Table 4. Summary description of the LMM fitting Vineland (general ability) and age as covariates. There were 48 subjects included in this analysis. The number of blocks played (Block number) and the interaction between the number of blocks had a significant effect.

#### Behavioural model

Second, we wanted to investigate whether behavioural characteristics (ADHD scoring) had an effect on game performance. Overall, 35 subjects were included in this analysis. Here we fitted block number, inattention, hyperactivity, age, and the interactions between block number and behavioural characteristics and number of blocks played as fixed factor, with age and subjects as random effects. Details of model’s results are presented in Table 5. The interaction terms fitted in this model (number of blocks, by hyperactivity, number of blocks by inattention, number of blocks by EFs) are shown in Figure 5.

|  | **Stimulus duration** | | |
| --- | --- | --- | --- |
| *Predictors* | *Estimates* | *CI* | *p* |
| (Intercept) | 1611.05 | 1351.56 – 1870.53 | **<0.001** |
| Block number | -118.95 | -162.01 – -75.88 | **<0.001** |
| Inattention | -44.21 | -459.46 – 371.04 | 0.836 |
| Hyperactivity | -51.76 | -386.17 – 282.65 | 0.764 |
| EF | 87.24 | -286.32 – 460.79 | 0.651 |
| Age | 41.99 | -512.06 – 596.04 | 0.883 |
| Block nr:Inattention | -69.30 | -148.38 – 9.77 | 0.086 |
| Block nr:Hyperactivity | -108.60 | -182.24 – -34.96 | **0.004** |
| Block nr:EF | -13.06 | -74.93 – 48.80 | 0.679 |
| **Random Effects** | | | |
| σ^2^ | 440152.81 | | |
| τ_00_ _subject_ | 214243.23 | | |
| τ_11_ _subject.age_months_ | 1295929.14 | | |
| ρ_01_ _subject_ | -0.08 | | |
| ICC | 0.72 | | |
| N _subject_ | 35 | | |
| Observations | 1563 | | |
| Marginal R^2^ / Conditional R^2^ | 0.029 / 0.727 | | |

Table 5. Model summary of the behavioural LMM, fitting behavioural characteristics as covariates. The number of blocks has a significant effect on reducing the stimulus duration. Among behavioural characteristics, there was a significant interaction between a higher number of blocks played and hyperactivity.

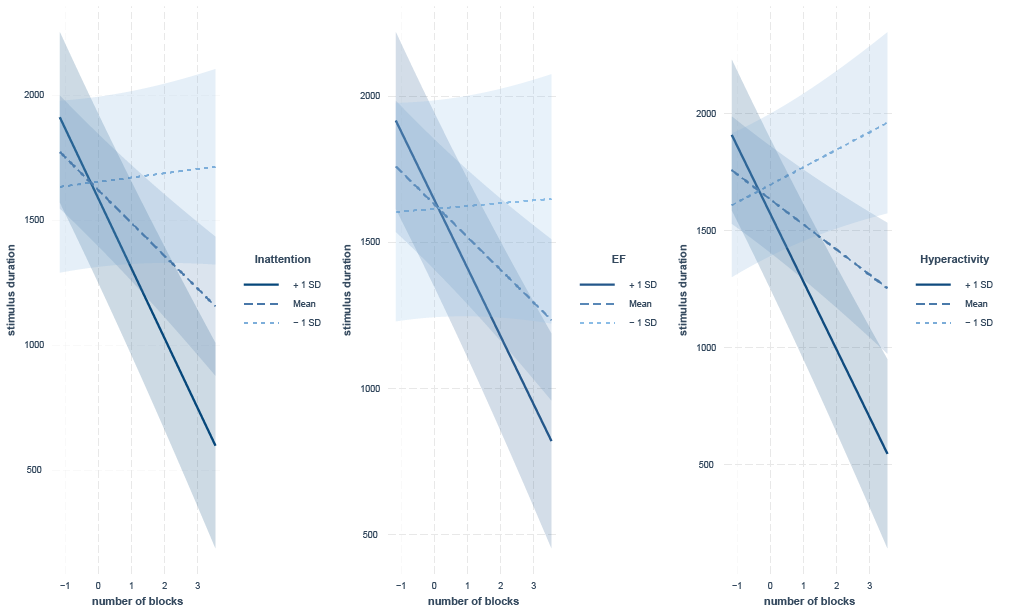

Figure 5. Here we show how different rates (mean ± 1 SD) of Inattention, EF, and Hyperactivity influenced game performance (stimulus duration) over time (number of blocks). While the interaction was only significant for Hyperactivity, the general trend is that increased behavioural (+1SD over the mean) symptoms exhibit a slower starting point (intercept), but a steeper learning curve over time. This case was particularly relevant for Hyperactivity.

### Study 4: Effects of genetic diagnosis on FarmApp performance and change over time

List of genes included in the FNG analysis:

Chromatin group= ARID1B, EHMT1, KAT6B, SETD5, SMARCA2

Synaptic= CASK, CTNNB1, DDX3X, DLG3, DYRK1A, PAK3, SHANK1, SHANK3, STXBP1, TRIO

Table 6 shows the LMM’s summary that explores change in performance over time, based on genetic diagnosis (FNG). As shown in the manuscript, there was a significant effect of the number of blocks played. This was particularly relevant for the *synaptic* genetic group.

|  | **Stimulus duration** | | |
| --- | --- | --- | --- |
| *Predictors* | *Estimates* | *CI* | *p* |
| (Intercept) | 1506.17 | 1065.44 – 1946.90 | **<0.001** |
| Block number | -113.99 | -176.28 – -51.69 | **<0.001** |
| Genetic group [Synaptic] | 562.05 | -137.77 – 1261.87 | 0.110 |
| Vineland | 368.81 | -261.06 – 998.69 | 0.221 |
| Age months | -159.91 | -594.04 – 274.22 | 0.458 |
| Block nr * genetic group [Synaptic] | -341.69 | -458.51 – -224.87 | **<0.001** |
| **Random Effects** | | | |
| σ^2^ | 446353.41 | | |
| τ_00_ _subject_ | 197272.04 | | |
| τ_11_ _subject.age_months_ | 698571.29 | | |
| ρ_01_ _subject_ | -0.30 | | |
| ICC | 0.71 | | |
| N _subject_ | 35 | | |
| Observations | 1419 | | |
| Marginal R^2^ / Conditional R^2^ | 0.132 / 0.749 | | |

Table 6. This table shows the output of the LMM fitting age, Vineland and block number, and interaction with FNG group, with subject and age as random effects. The output shows there was a significant effect of block number, and a significant interaction with group, where the other group showed an increase improvement in the sheep game (lower stimulus duration).
